# Evidence of Epistatic Interactive Effects of HK1 and GCK Genes on Circulating Hemoglobin A1c Levels

**DOI:** 10.64898/2026.05.01.26352221

**Authors:** Lihua Wang, Joseph H. Lee, Bharat Thyagarajan, Anatoliy I. Yashin, E. Warwick Daw, Thomas T. Perls, Kaare Christensen, Joseph M. Zmuda, Michael A. Province, Ping An

## Abstract

**Background:** Hemoglobin A1c (HbA1c), an important diagnostic biomarker for type 2 diabetes (T2D), is also associated with aging, cognitive performance, and mortality. To identify epistatic interactions, we assessed 133 known gene variants associated with HbA1c among 3,778 non-diabetic subjects of European ancestry in the Long Life Family Study (LLFS).

**Methods:** We applied Bayesian Imputation Based Association Mapping (BIMBAM) to identify significant pairwise epistatic interactions among genetic variants that were previously shown to be associated with levels of HbA1c. To take into account confounding effects, we adjusted age, sex, field centers, body mass index (BMI), and genetic principal components (PCs).

**Results:** This analysis yielded seven pairs with log_10_(BF)>10; of those, six pairs were confirmed using a full-term mixed regression model. Specifically, these included significant interactions of *HK1-rs17476364* with variants in *GCK* (rs2971670, rs4607517) or *G6PC2* (rs560887), as well as between *HK1-rs16926246* and the same variants (*P* values for each term ≤ 7.14×10^-3^). All epistatic interactions between *HK1* and *GCK*, and between *HK1* and *G6PC2* were replicated in two large independent studies (namely, Framingham Offspring Study, *P* < 0.05; Health and Retirement Study, *P* < 0.05).

**Conclusion:** The present study revealed that *HK1* and *GCK* interact to contribute to regulating levels of HbA1c and are likely to be involved in molecular mechanisms underlying healthy aging processes.

## 1 Introduction

HbA1c is formed by nonenzymatic glycation of hemoglobin and has been widely used as a diagnostic biomarker of type 2 diabetes (T2D). In addition to T2D, HbA1c is associated with various metabolic aging and cardiovascular aging in diabetic and non-diabetic adults[1]. Moreover, HbA1c had a causal effect on white matter brain aging with higher level showing accelerated brain aging [2]. Higher HbA1c (particularly > 8.5%) was associated with substantially increased all-cause mortality [3]. Therefore, genetic regulation of HbA1c might shed light not only on the molecular mechanisms of HbA1c regulation, but also on the molecular pathogenesis of aging. The estimated heritability of HbA1c is 40%-60% [4, 5]. However, the identified GWAS loci explained only ∼6% of the heritability [6]. The remaining unexplained heritability is often referred to as ‘missing heritability’, and gene-gene interactions (i.e. epistatic interaction) may contribute to the missing heritability.

HbA1c is not regulated by individual genes in isolation, but rather is controlled by coordinated interactions within the gene network. The commonly adopted GWAS approach only estimates the effects of individual genetic variants on HbA1c, but overlooks joint modelling their collective effects of multiple genes in the pathways involved. As a result, Polygenic Risk Scores (PRS) - based on identified variants from GWAS capture only linear additive effects - explain just 3%-10% of HbA1c variance [7]. We posit that epistatic SNP-SNP interactions may reveal additional genetic contribution toward HbA1c and improve predictive power of PRS. Since even pairwise epistatic analyses at the genome-wide level is computationally demanding and memory intensive, we conducted a hypothesis-driven analysis here restricted to interactions among genetic variants that were previously reported to be associated with HbA1c. For this purpose, we used the data from the baseline visit for 3,778 non-diabetic participants of European descendants from the LLFS. We initially considered 143 candidate SNPs, of which 133 were available in LLFS and included in the analysis.

## 2 Methods

### 2.1 LLFS

#### Subjects

We performed initial epistatic analysis utilizing 3,778 non-diabetic European participants from the LLFS baseline visit. Diabetes status was defined as self-reported diabetes, and/or use of diabetes-relevant medications. LLFS is a multinational and multigenerational longitudinal family-based study. To investigate the factors related to exceptional healthy aging, LLFS recruited exceptional long-lived pedigrees from four field centers including Boston University Medical Center in Boston (BU), Columbia University Irving Medical Center in New York (NY), the University of Pittsburgh in Pittsburgh (PT), and the University of Southern Denmark field center (DK).

#### HbA1c Assay

HbA1c in LLFS was measured in EDTA whole blood using an automated high-performance liquid chromatography (HPLC) method on the Tosoh HPLC Glycohemoglobin Analyzer (Tosoh Medics, Inc., San Francisco, CA 94080) at the University of Minnesota Advanced Research and Diagnostics Laboratory. This method was calibrated to standard values established by the National Glycohemoglobin Standardization Program (NGSP). The coefficient of variation for HbA1c measurement ranged from 1.4% to 1.9%.

#### Sequencing

Genotypes were obtained from whole genome sequencing data (WGS) using 150-bp paired-end reads on an Illumina sequencer at the McDonnell Genome Institute (MGI), Washington University in Saint Louis. Whole genome sequencing reads were processed in two main steps. The first step, including read alignment and post-alignment quality control, was conducted by MGI. Briefly, reads were aligned to Genome Reference Consortium Human Build 38 (GRCh38) using the Burrows-Wheeler Aligner (BWA-MEM 0.7.15). Duplicated reads were marked with Picard 2.4.1, base quality score was recalibrated using GATK BaseRecalibrator 3.6, and the aligned reads were converted to CRAM format using SAMtools 1.3.1. The second step, consisting of variant calling and post-calling quality control was carried out at the Division of Statistical Genomics, Washington University in St. Louis [8]. In brief, variant calling was performed by using GATK HaplotypeCaller on the CRAM files to obtain subject-level gVCF files, which were then combined with GATK HaplotypeCaller, followed by joint genotyping with GATK GenotypeGVCFs. Post-calling quality control excluded low-quality samples with estimated contamination (FREEMIX > 0.03), low mean sequencing depth (< 20x), or excessive Mendelian errors identified using LOKI 2.4.5 [9]. We also removed low quality genetic variants based on abnormal sequencing depth (<20X or >300x), significant deviation from Hardy-Weinberg equilibrium (HWE; p<1e-6), or elevated heterozygosity (>0.55). In addition, a small number of sample swaps were identified and corrected by comparing WGS and GWAS genotypes and by comparing the genotypes of family members.

### 2.2 Framingham Heart Study Offspring Cohort (FOS)

Diabetes status was defined as self-reported diabetes, or use of diabetes-relevant medications. FOS participants with diabetes were excluded. Non-diabetic participants from FOS examination 7 data (phs000007.v35.pht000156.v7.p11.c1.hba1_7s; n=1456; average age: 60.49 ± 0.25) were used to replicate the identified epistatic interactions. The Framingham Heart Study is a longitudinal multigenerational cohort designed to identify common risk factors for cardiovascular disease [10, 11]. In the FOS, HbA1c levels were measured using high-performance liquid chromatography after an overnight dialysis against normal saline to remove the labile fraction [12]. All assays were standardized to the Diabetes Control and Complications Trial reference method.

As part of the National Heart, Lung and Blood Institute’s TOPMed phase I, WGS genotypes from FOS (phg002135.v1.TOPMed_WGS_Framingham_v6_frz10_SNPs.genotype-calls-vcf) were sequenced at >30× depth of coverage from the Broad Institute of the Massachusetts Institute of Technology and Harvard. The joint calling of all samples, along with quality control of variants and samples were performed by the TOPMed Informatics Resource Center at University of Michigan [13].

### 2.3 Health and Retirement Study (HRS)

Diabetes status was defined as self-reported diabetes, or use of diabetes-relevant medications. HRS participants with diabetes were excluded. Non-diabetic participants from HRS (phs000428.v2.p2) (n=2094; average age: 73.55 ± 0.21) were also used to further replicate the identified epistatic interactions. HRS is a longitudinal study of noninstitutionalized adults aged ≥ 50 years in the United States [14]. HbA1c levels in 2014 HRS biomarker data were measured by dried blood spots. The HbA1c levels were adjusted to be consistent with National Health and Nutrition Examination Survey (NHANES)[15].

DNA for HRS participants was extracted from saliva samples using Oragene^TM^ saliva kits (DNA Genotek, Ontario, Canada). The extracted DNA was subsequently genotyped on the Illumina HumanOmni2.5 array using the calling algorithm GenomeStudio version 2011.1, Genotyping Module version 1.9.4 and GenTrain version 1.0 at the Center for Inherited Disease Research (CIDR). The SNP annotation from CIDR (“HumanOmni2.5-8v1_C”) used genome build 37/hg 19. Quality control of the genotypes was conducted by the Genetics Coordinating Center at the University of Washington.

### 2.4 Statistical analysis

#### Interaction analyses in LLFS

A total of 143 known SNPs were reported for HbA1c [6, 16], of which, 10 (four indels, four multi-allelic variants, and two in complex variant regions) were absent, and 133 were present in final cleaned LLFS WGS data. HbA1c levels for 3,778 non-diabetic LLFS participants at visit 1 were adjusted by age, age squared, sex, field centers, BMI, and 10 genetic principal components. BMI was include as a covariate to estimate genetic interaction on HbA1c independent of adiposity. The single-SNP and two-SNP effects of 133 SNPs on adjusted HbA1c residuals were screened using Bayesian imputation-based association mapping (BIMBAM) [17]. *P* values for both single-SNP and two-SNP effect were based on 10000 permutations. The significance was determined based on log_10_(BF) > 3 for single-SNP (133 SNPs), and log_10_(BF)>10 for two-SNP test (8778 SNP pairs). The cutoff of log_10_(BF) > 3 was used to indicate strong evidence for association. To account for the substantially large model space and increased multiple testing burden in two-SNP test, a more stringent threshold log_10_(BF)>10 was adopted. Since BIMBAM does not account for relatedness among family members, the significance of interaction between the identified SNP pairs were re-evaluated using a mixed-effects model in SAS 9.4 (SAS Institute Inc., Cary, NC.) to account for non-independence among family members. Concordance between BIMBAM results and the mixed-effects model provides reassurance about robustness of our findings.

To examine whether interplay between the identified significant SNP pairs regulate glycemic control or red blood cell (RBC) turn over, we further adjusted fasting glucose (n=3,339) and Hemoglobin (Hb; n=3,339) with the same covariates as for HbA1c, and additionally assessed the interaction effects of these SNP pairs on generated glucose residuals and Hb residuals.

#### Replication in FOS

For the detected significant interactions, the SNPs were extracted from FOS WGS data. The downloaded FOS dataset lacked genetic principal components, limiting our ability to control for population structure. Therefore, HbA1c levels of 1,456 non-diabetic FOS participants were only adjusted by age, age squared, sex, and BMI. *P* values for interaction effect of the identified SNP pairs on HbA1c residuals were estimated using linear model in SAS 9.4 (SAS Institute Inc., Cary, NC.).

#### Replication in HRS

The significant interactions were also re-examined using HRS data. Two SNPs (*HK1-rs17476364* and *GCK-rs2971670*) were not present in HRS genotype data. For these two SNPs, two proxy SNPs including *HK1-rs16926246* (r^2^=0.7 with *HK1-rs17476364*), and *GCK-rs2971667* (r^2^=0.98 with *GCK-rs2971670*) were used in our replication. Similarly, HbA1c levels of 2,094 non-diabetic HRS participants were adjusted by age, age squared, sex, BMI and 10 genetic principal components. *P* values for interaction effect of identified SNP pairs on HbA1c residuals were estimated with linear model using SAS 9.4 (SAS Institute Inc., Cary, NC.).

## 3 Results

### 3.1 Characteristics of Samples

As shown in Table 1, a total of 3,778 non-diabetic LLFS participants including 1,037 from BU, 1,014 from DK, 767 from NY, and 960 from PT was included in this study. About ∼56% of these samples (53.35%-57.08% by field centers) were female. The average age of all samples was 69.81 years. NY (73.00±0.59) and PT participants (71.03±0.52) are significantly older than BU (68.69±0.49) and DK participants (67.41±0.44), showing P value < 0.0001. Among the four field centers, participants from DK had the lowest average HbA1c levels (*P*<0.01 vs BU; *P*<0.0001 vs NY; *P*<0.0001 vs PT).

**Table 1.**
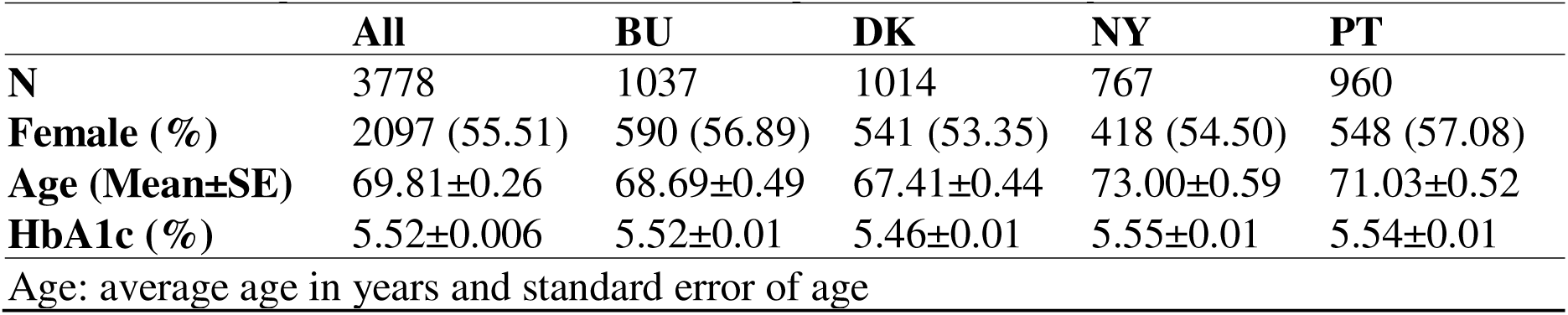
Descriptive statistics for LLFS samples used in analyses.

| **Table 1. Descriptive statistics for LLFS samples used in analyses** |  |  |  |  |  |
| --- | --- | --- | --- | --- | --- |
|  | **All** | **BU** | **DK** | **NY** | **PT** |
| **N** | 3778 | 1037 | 1014 | 767 | 960 |
| **Female (%)** | 2097 (55.51) | 590 (56.89) | 541 (53.35) | 418 (54.50) | 548 (57.08) |
| **Age (Mean±SE)** | 69.81±0.26 | 68.69±0.49 | 67.41±0.44 | 73.00±0.59 | 71.03±0.52 |
| **HbA1c (%)** | 5.52±0.006 | 5.52±0.01 | 5.46±0.01 | 5.55±0.01 | 5.54±0.01 |
| Age: average age in years and standard error of age |  |  |  |  |  |

### 3.2 SNP interactions regulating HbA1c

As shown in Table 2 (BIMBAM) and Supplemental Table 1 (Linear mixed model), single-SNP test identified five significant SNPs (*HK1*-rs17476364, *HK1*-rs16926246, *GCK*-rs2971670, *GCK*-rs4607517, and *G6PC2*-rs560887; r^2^=0.71 between two *HK1* SNPs; r^2^=0.98 between two *GCK* SNPs) in LLFS. All five SNPs are intronic variants, each showing significant association with HbA1c (log_10_(BF) > 3; *P*<1×10^-3^). The top two SNPs resided within HK1 (*HK1*-rs17476364: *P*=1.00×10^-11^, log_10_(BF)=9.376 ; and *HK1*-rs16926246: *P*=7.02×10^-9^, log_10_(BF)=7.494). The risk allele was defined as the minor allele. C allele of *HK1*-rs17476364 and T allele of *HK1*-rs16926246 were associated with lower HbA1c levels (Fig 1A). Two-SNP test results were also given in Table 2. Seven significant SNP pairs were nominated by two-SNP BIMBAM test (Table 2: log_10_(BF)=12.992 for *HK1*-rs17476364 and *GCK*-rs2971670; log_10_(BF)=12.883 for *HK1*-rs17476364 and *G6PC2*-rs560887; log_10_(BF)=12.851 for *HK1*-rs17476364 and *GCK*-rs4607517; log_10_(BF)=11.197 for *HK1*-rs16926246 and *GCK*-rs2971670; log_10_(BF)=11.068 for *HK1*-rs16926246 and *GCK*-rs4607517; log_10_(BF)=10.877 for *HK1*-rs16926246 and *G6PC2*-rs560887; and log_10_(BF)=10.154 for *HK1*-rs17476364 and *HK1*-rs16926246).

**Figure 1.**
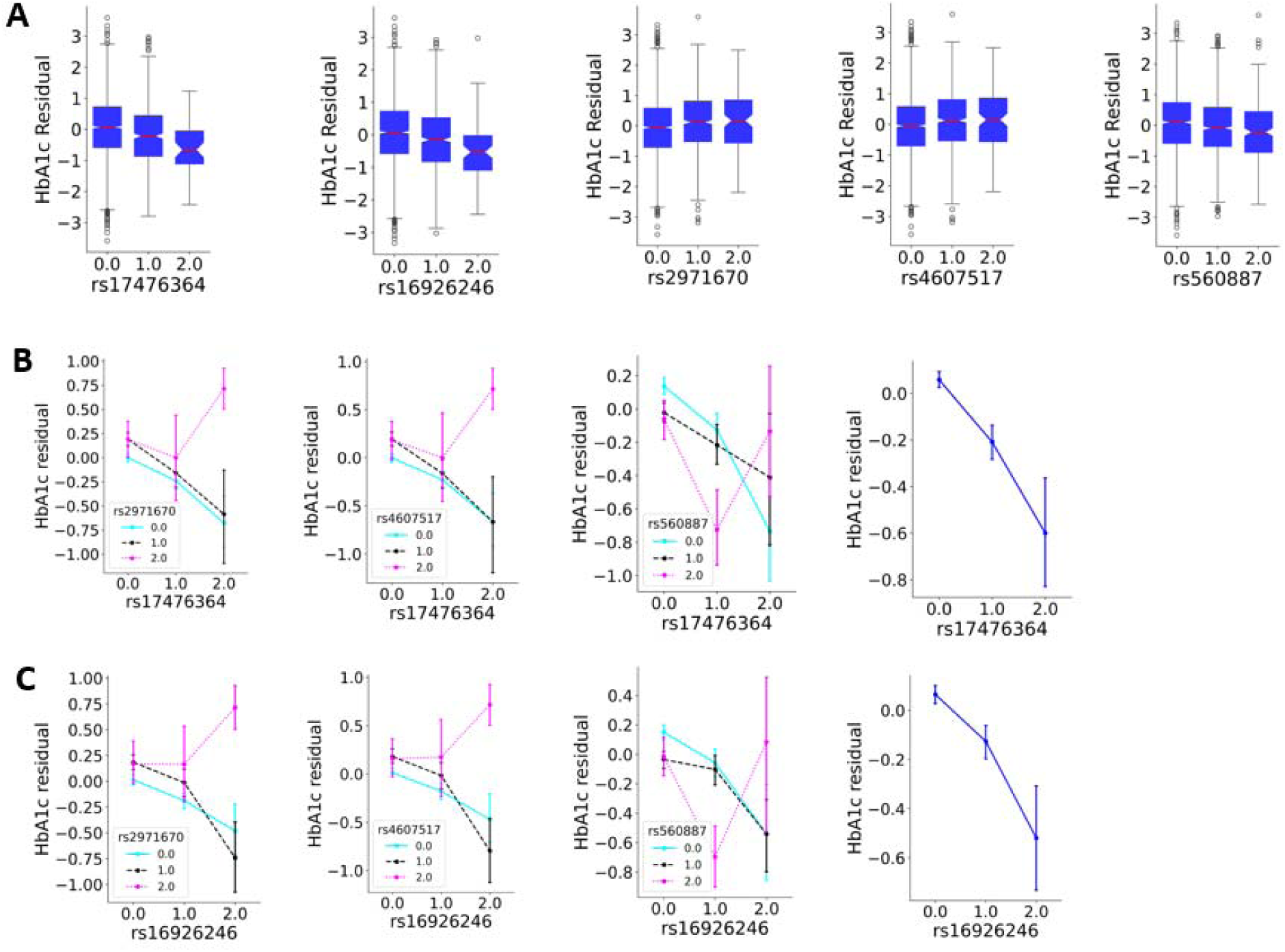
Box plots of HbA1c residuals and plots of interaction effect among six pair SNPs on HbA1c residuals. A) Box plots of HbA1c residuals for five SNPs (rs17476364, rs16926246, rs2971670, rs4607517, and rs560887). The x-axis displayed the dosage of SNPs (number of risk allele). The boxes (blue color) bound the IQR, and the lines in red depicted the median of HbA1c residuals. Tukey-style whiskers extend to 1.5 × IQR beyond the box. Genotype coding 0/1/2 indicates the number of excluded.. B) Plots of interaction effect among rs17476364 and three SNPs on HbA1c residuals. The x-axis displayed the dosage of rs17476364 (number of risk allele C). The association of rs17476364 and HbA1c residuals was colored blue in dosage of 0 of second SNP, black in dosage of 1 of second SNP, and magenta in dosage of 2 of second SNP. Genotype coding 0/1/2 indicates the number of minor alleles. C) Plots of interaction effect among rs16926246 and three SNPs on HbA1c residuals. The x-axis displayed the dosage of rs16926246 (number of risk allele C). The association of rs16926246 and HbA1c residuals was colored blue in dosage of 0 of second SNP, black in dosage of 1 of second SNP, and magenta in dosage of 2 of second SNP. Genotype coding 0/1/2 indicates the number of minor alleles.

**Table 2.** Association results of best single SNP and SNP pairs for HbA1c in LLFS.

| rsID | Chr | Position | Genes | RAF | Log10(BF) | Rank | P |
| --- | --- | --- | --- | --- | --- | --- | --- |
| rs17476364 | 10 | 69334748 | *HK1(intron)* | C: 0.10 | 9.376 | 1 | $1.00 \times 10^{-4}$ |
| rs16926246 | 10 | 69333636 | *HK1(intron)* | T: 0.14 | 7.494 | 2 | $1.00 \times 10^{-4}$ |
| rs2971670 | 7 | 44186502 | *GCK (intron)* | T: 0.17 | 3.516 | 3 | $1.00 \times 10^{-4}$ |
| rs4607517 | 7 | 44196069 | *GCK (intron)* | A: 0.17 | 3.366 | 4 | $1.00 \times 10^{-4}$ |
| rs560887 | 2 | 168906638 | *G6PC2 (intron)* | T: 0.28 | 3.226 | 5 | $1.00 \times 10^{-4}$ |
| SNP pair 1 (1,3) |  |  |  |  | 12.992 | 1,3 |  |
| SNP pair 2 (1,5) |  |  |  |  | 12.883 | 1,5 |  |
| SNP pair 3 (1,4) |  |  |  |  | 12.851 | 1,4 |  |
| SNP pair 4 (2,3) |  |  |  |  | 11.197 | 2,3 |  |
| SNP pair 5 (2,4) |  |  |  |  | 11.068 | 2,4 |  |
| SNP pair 6 (2,5) |  |  |  |  | 10.877 | 2,5 |  |
| SNP pair 7 (1,2) |  |  |  |  | 10.154 | 1,2 |  |

RAF: risk allele and frequency; the risk allele was defined as the minor allele
P: permutation-based p-value for single-SNP association tests using BIMBAM

Since significant two-SNP BIMBAM tests only indicated that two-SNP model (with or without interactions) better explained variation in HbA1c levels than the single-SNP model, we utilized the same LLFS samples and further examined the interactions among seven SNP pairs using mixed effect model implemented in SAS (Table 3). The corresponding genotype counts for five SNPs are shown in Supplemental Table 2. Except for one SNP pair (*HK1*-rs17476364 and *HK1*-rs16926246), significant interactions were identified between the remaining six SNP pairs that had *P*< 7.14×10^-3^ (Bonferroni-corrected for 7 tests: 0.05/7). These six pairs include HK1-rs17476364 and GCK-rs2971670 (*P*=2.47×10^-3^, R^2^=0.187%), HK1-rs17476364 and GCK-rs4607517 (*P*=2.37×10^-5^ , R^2^=0.189%), HK1-rs17476364 and G6PC2-rs560887 (*P*=1.00×10^-3^ , R^2^=0.201%), HK1-rs16926246 and GCK-rs2971670 (*P*=1.33×10^-4^ , R^2^=0.311%), HK1-rs16926246 and GCK-rs4607517 (*P*=9.50×10^-5^ , R^2^=0.323%), and HK1-rs16926246 and G6PC2-rs560887 (*P*=3.43×10^-5^ , R^2^=0.198%). The interactions between six SNP pairs were clearly illustrated in Fig 1B and Fig 1C. When analyzed independently, both *HK1-rs17476364* and *HK1-rs16926246*, showed a dose-dependent association with HbA1c levels, where one risk allele corresponded to lower HbA1c than none, and two risk alleles to the lowest HbA1c. Upon stratification by the second SNP, however, this pattern was reversed in individuals homozygous for its risk allele, displaying a ‘V’-shaped relationship with the lowest HbA1c observed in those homozygous for the primary SNP and heterozygous for the second SNP. The association pattern remained unchanged in the other genotype groups of the second SNP.

**Table 3.** Confirmed interaction of SNP pairs for HbA1c in LLFS.

| **Model** | **Effect** | **N** | **HbA1c Pval\*** |
| --- | --- | --- | --- |
| **1** | *HK1*-rs17476364 | 3402 | $4.40 \times 10^{-4}$ |
| | *GCK*-rs2971670 | 3402 | $2.47 \times 10^{-6}$ |
| | *HK1* -rs17476364* *GCK* -rs2971670 | 3402 | $2.47 \times 10^{-3}$ |
| **2** | *HK1*-rs17476364 | 3382 | $6.28 \times 10^{-4}$ |
| | *GCK*-rs4607517 | 3382 | $2.55 \times 10^{-8}$ |
| | *HK1* -rs17476364* *GCK* -rs4607517 | 3382 | $2.37 \times 10^{-5}$ |
| **3** | *HK1*-rs17476364 | 3345 | $7.21 \times 10^{-10}$ |
| | *G6PC2*-rs560887 | 3345 | $9.14 \times 10^{-1}$ |
| | *HK1* -rs17476364* *G6PC2* -rs560887 | 3345 | $1.00 \times 10^{-3}$ |
| **4** | *HK1*-rs16926246 | 3272 | $5.16 \times 10^{-3}$ |
| | *GCK*-rs2971670 | 3272 | $2.26 \times 10^{-8}$ |
| | *HK1* -rs16926246* *GCK* -rs2971670 | 3272 | $1.33 \times 10^{-4}$ |
| **5** | *HK1*-rs16926246 | 3288 | $4.18 \times 10^{-3}$ |
| | *GCK*-rs4607517 | 3288 | $3.62 \times 10^{-8}$ |
| | *HK1* -rs16926246* *GCK* -rs4607517 | 3288 | $9.50 \times 10^{-5}$ |
| **6** | *HK1*-rs16926246 | 3238 | $7.81 \times 10^{-9}$ |
| | *G6PC2*-rs560887 | 3238 | $4.38 \times 10^{-1}$ |
| | *HK1* -rs16926246* *G6PC2* -rs560887 | 3238 | $3.43 \times 10^{-5}$ |
| **7** | *HK1*-rs17476364 | 3602 | $7.00 \times 10^{-3}$ |
| | *HK1*-rs16926246 | 3602 | $9.47 \times 10^{-2}$ |
|  | *HK1* -rs17476364* *HK1* -rs16926246 | 3602 | . |
| **\*: P value derived from a mixed model using categorical SNP genotype data** |  |  |  |

We further assessed the interaction effects between six SNP pairs using two independent external studies: FOS (n=1,456) and HRS (n=2,094). As shown in Supplemental table 2, in FOS, all but two SNP pairs (*HK1-rs16926246* and *GCK-rs2971670*, and *HK1-rs16926246* and *GCK-rs4607517*) showed significant interaction effects (*P*<0.05).

Because *HK1-rs17476364* and *GCK-rs2971670* were not genotyped in HRS, we selected their proxy SNPs rs10159477 (r^2^=0.7 with *HK1-rs17476364*) and rs2971667 (r^2^=0.98 with *GCK-rs2971670*), and validated the interaction effects between four SNP pairs (rs10159477 and rs2971667: *P*=0.0188; rs10159477 and rs4607517: *P*=0.0185; rs16926246 and rs2971670: *P*=0.0462; rs16926246 and rs4607517: *P*=0.0418; Supplemental Table 3). Overall, all SNP pair (*HK1-rs16926246* and*GCK-rs2971670*) reached *P*<0.05 in FOS and/or HRS. Notably, two *HK1*-*GCK* SNP pairs (*HK1-rs17476364* and*GCK-rs2971670*; *HK1*-rs17476364 and*GCK*-rs4607517) reached nominal significance in both FOS and HRS. In addition, two *HK1*-*G6PC2* SNP pairs (*HK1-rs17476364* and *G6PC2-rs560887*; *HK1-rs16926246* and *G6PC2-rs560887*) passed the Bonferroni corrected threshold (*P*<8.33×10^-3^, 0.05/6) in FOS.

### 3.3 SNP interactions regulating Hb and Glucose

Because HbA1c is generated through non-enzymatic glycation of Hb, its level is affected by both fasting blood glucose and the concentration of Hb in the blood. Of the five SNPs examined (genotype counts in Supplemental Table 4), two (*HK1*-rs17476364 and *HK1*-rs16926246; Supplemental Table 1; Fig 2A) were significantly associated with Hb and showed effects opposite in direction to those observed for HbA1c. In contrast, the remaining three (*GCK*-rs2971670, *GCK*-rs4607517, and *G6PC2*-rs560887; Supplemental Table 1; Fig 3A) were significantly associated with glucose and exhibited effect directions consistent with those for HbA1c.

**Figure 2.**
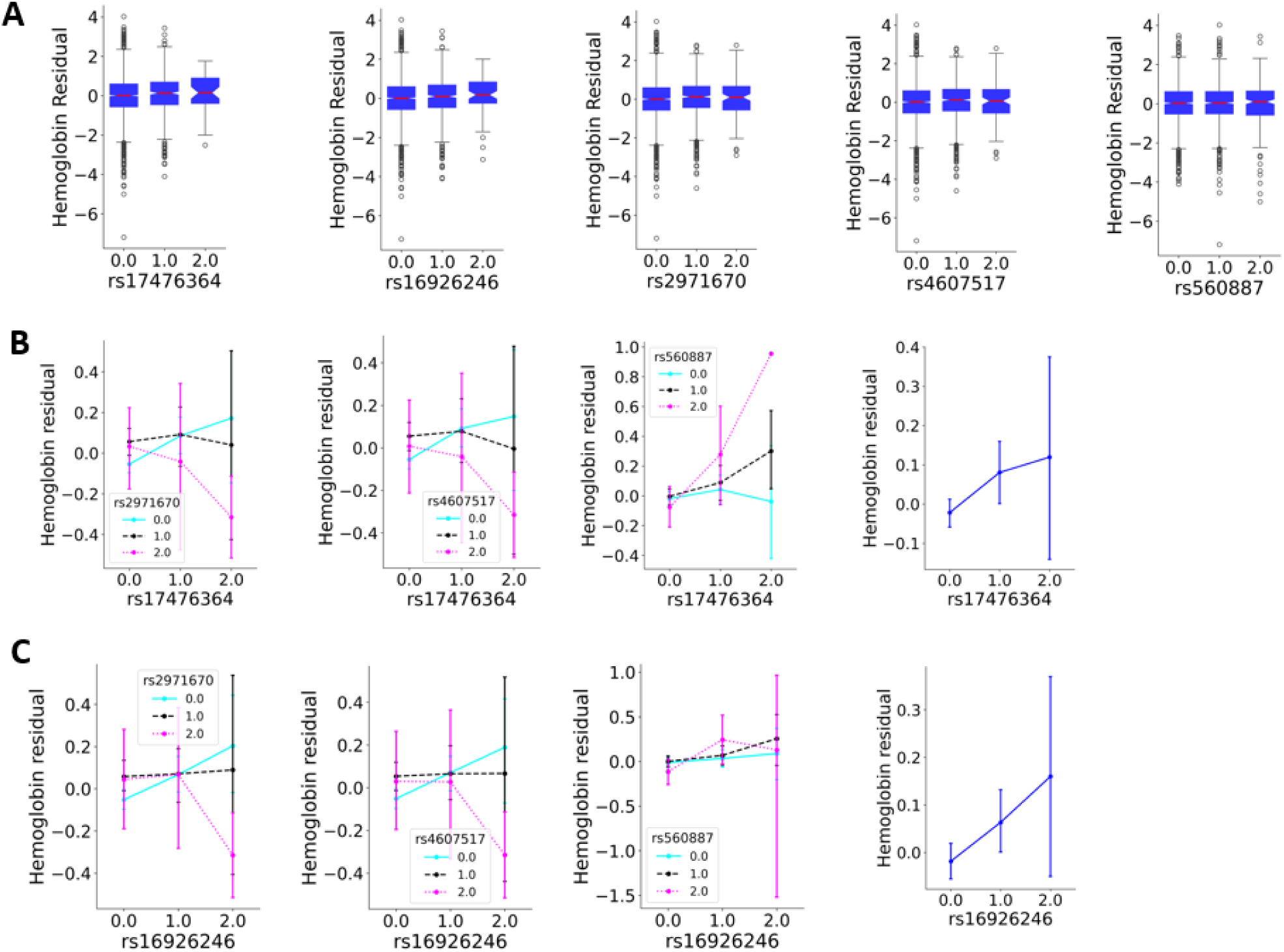
Box plots of Hemoglobin residuals and plots of interaction effect among six pair SNPs on Hemoglobin residuals. A) Box plots of Hemoglobin residuals for five SNPs (rs17476364, rs16926246, rs2971670, rs4607517, and rs560887). The x-axis displayed the dosage of SNPs (number of risk allele). The boxes (blue color) bound the IQR, and the lines in red depicted the median of Hemoglobin residuals. Tukey-style whiskers extend to 1.5 × IQR beyond the box. B) Plots of interaction effect among rs17476364 and three SNPs on Hemoglobin residuals. The x-axis displayed the dosage of rs17476364 (number of risk allele C). The association of rs17476364 and Hemoglobin residuals was colored blue in dosage of 0 of second SNP, black in dosage of 1 of second SNP, and magenta in dosage of 2 of second SNP. C) Plots of interaction effect among rs16926246 and three SNPs on Hemoglobin residuals. The x-axis displayed the dosage of rs16926246 (number of risk allele C). The association of rs16926246 and Hemoglobin residuals was colored blue in dosage of 0 of second SNP, black in dosage of 1 of second SNP, and magenta in dosage of 2 of second SNP.

**Figure 3.**
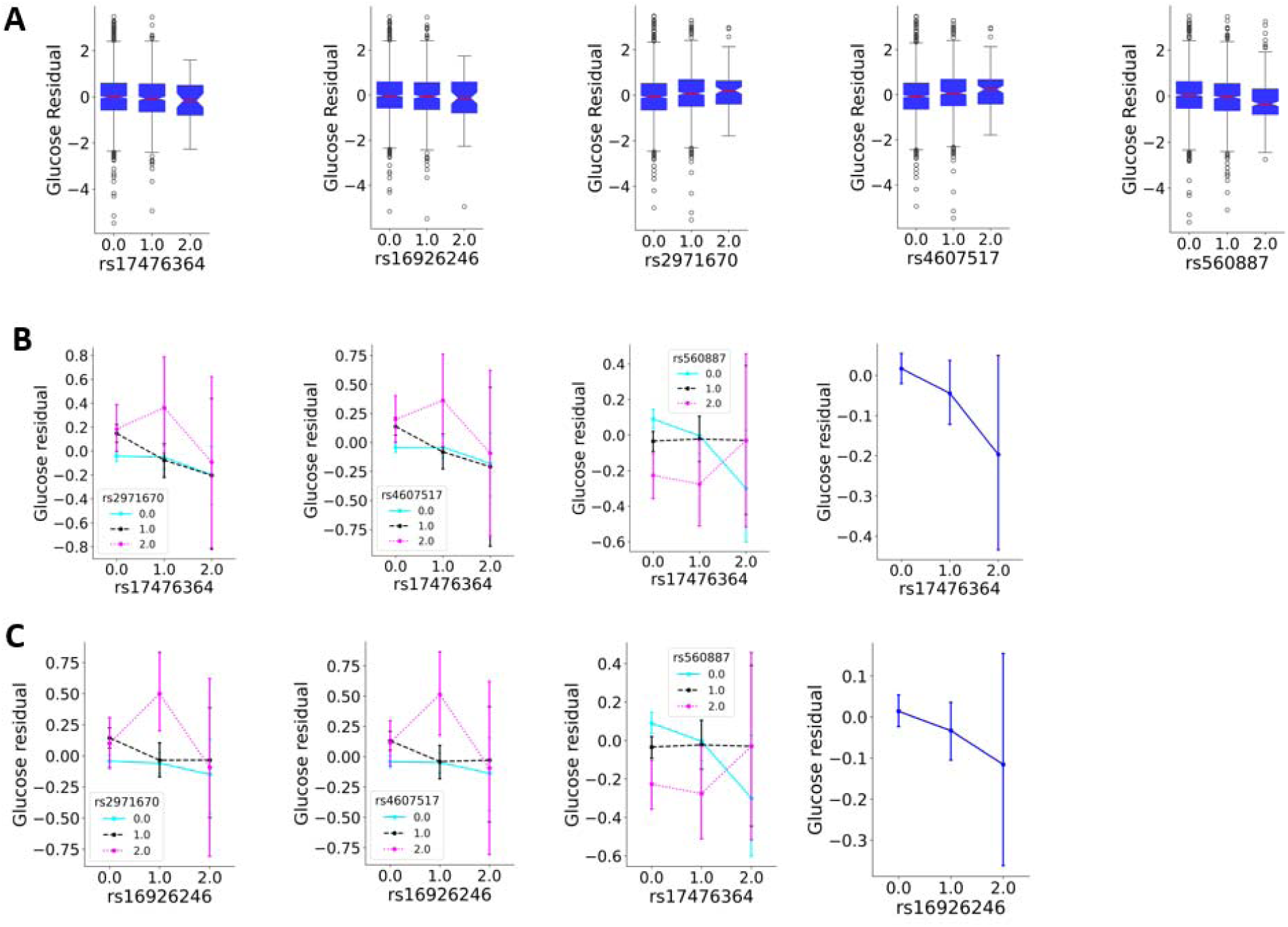
Box plots of Glucose residuals and plots of interaction effect among six pair SNPs on Glucose residuals. A) Box plots of Glucose residuals for five SNPs (rs17476364, rs16926246, rs2971670, rs4607517, and rs560887). The x-axis displayed the dosage of SNPs (number of risk allele). The boxes (blue color) bound the IQR, and the lines in red depicted the median of Glucose residuals. Tukey-style whiskers extend to 1.5 × IQR beyond the box. B) Plots of interaction effect among rs17476364 and three SNPs on Glucose residuals. The x-axis displayed the dosage of rs17476364 (number of risk allele C). The association of rs17476364 and Glucose residuals was colored blue in dosage of 0 of second SNP, black in dosage of 1 of second SNP, and magenta in dosage of 2 of second SNP. C) Plots of interaction effect among rs16926246 and three SNPs on Glucose residuals. The x-axis displayed the dosage of rs16926246 (number of risk allele C). The association of rs16926246 and Glucose residuals was colored blue in dosage of 0 of second SNP, black in dosage of 1 of second SNP, and magenta in dosage of 2 of second SNP.

To examine whether the interaction effects of the six SNP pairs modulate glucose or Hb, we assessed their interaction effects on both Hb concentrations and glucose level. As shown in Table 4, Fig 2 and Fig 3, except for one snp pair (*HK1-rs16926246* and *G6PC2-rs560887*), five SNP pairs demonstrated significant interactions on Hb (Pval<0.05), whereas none of the six pairs showed interaction effects on glucose. The interaction effect on Hb were illustrated on Fig 2B and Fig 2C. When examined individually, *HK1-rs17476364* and *HK1-rs16926246*, each exhibited a dose-dependent association with Hb, where increasing numbers of risk alleles corresponded to progressively higher Hb levels. However, this relationship was reversed after stratification by a second SNP—specifically in individuals homozygous for the risk allele of *GCK*-rs2971670 or *GCK*-rs4607517, or homozygous for the alternate allele of *G6PC2*-rs560887. The lowest Hb levels were observed in individuals who were double homozygous for the risk allele of four SNP pairs: *HK1-rs17476364* and *GCK*-rs2971670, *HK1-rs17476364* and *GCK*-rs4607517, *HK1-rs16926246* and *GCK*-rs2971670, and *HK1-rs16926246* and *GCK*-rs4607517.

**Table 4.** Association results of SNP pairs with HbA1c, glucose and Hb.

| Model | Effect | N | Pval* |  |
| --- | --- | --- | --- | --- |
|  |  |  | Glucose | Hb |
| 1 | HK1-rs17476364 | 3309 | $3.08 \times 10^{-1}$ | $4.62 \times 10^{-1}$ |
| | GCK-rs2971670 | 3309 | $3.96 \times 10^{-1}$ | $4.90 \times 10^{-3}$ |
| | HK1-rs17476364*GCK-rs2971670 | 3309 | $2.79 \times 10^{-1}$ | $3.39 \times 10^{-2}$ |
| 2 | HK1-rs17476364 | 3285 | $3.48 \times 10^{-1}$ | $4.35 \times 10^{-1}$ |
| | GCK-rs4607517 | 3285 | $4.70 \times 10^{-1}$ | $4.78 \times 10^{-3}$ |
| | HK1-rs17476364*GCK-rs4607517 | 3285 | $2.45 \times 10^{-1}$ | $3.46 \times 10^{-2}$ |
| 3 | HK1-rs17476364 | 3253 | $7.35 \times 10^{-1}$ | $2.36 \times 10^{-6}$ |
| | G6PC2-rs560887 | 3253 | $5.80 \times 10^{-1}$ | $1.76 \times 10^{-3}$ |
| | HK1-rs17476364*G6PC2-rs560887 | 3253 | $3.05 \times 10^{-1}$ | $1.10 \times 10^{-5}$ |
| 4 | HK1-rs16926246 | 3196 | $4.81 \times 10^{-1}$ | $2.21 \times 10^{-1}$ |
| | GCK-rs2971670 | 3196 | $2.11 \times 10^{-1}$ | $1.84 \times 10^{-2}$ |
| | HK1-rs16926246*GCK-rs2971670 | 3196 | $1.67 \times 10^{-1}$ | $1.11 \times 10^{-2}$ |
| 5 | HK1-rs16926246 | 3176 | $4.78 \times 10^{-1}$ | $2.43 \times 10^{-1}$ |
| | GCK-rs4607517 | 3176 | $3.24 \times 10^{-1}$ | $1.17 \times 10^{-2}$ |
| | HK1-rs16926246*GCK-rs4607517 | 3176 | $1.92 \times 10^{-1}$ | $1.20 \times 10^{-2}$ |
| 6 | HK1-rs16926246 | 3147 | $4.74 \times 10^{-1}$ | $2.93 \times 10^{-4}$ |
| | G6PC2-rs560887 | 3147 | $5.92 \times 10^{-2}$ | $4.00 \times 10^{-1}$ |
| | HK1-rs16926246*G6PC2-rs560887 | 3147 | $9.67 \times 10^{-1}$ | $2.74 \times 10^{-1}$ |

**Pval\*: P value derived from a mixed model using categorical SNP genotype data**

## 4 Discussion

Using 3,778 nondiabetic European LLFS Participants, we assessed pairwise epistatic genetic regulation of HbA1c using 133 known HbA1c-associated SNPs. Six significant SNP-SNP interactions between *HK1* and *GCK* genes, and between *HK1* and *G6PC2* genes were detected. Interaction effects on HbA1c between all *HK1-GCK* and *HK1-G6PC2* pairs were independently replicated in FOS or/and HRS (*P*<0.05). More interestingly, except for one SNP pair (*HK1-rs16926246* and *G6PC2-rs560887*), we also observed significant interaction effects among the remaining *HK1-GCK* and *HK1-G6PC2* SNP pairs on regulation of Hb levels. Variance explained by these interaction terms is small (∼0.18-0.32%), which is typical for interaction terms in complex traits. The strength of these findings lies in consistent replication and biological coherence rather than variance magnitude.

HbA1c is produced by non-enzymatical glycation of hemoglobin within red blood cells. Therefore, genetic variants can influence HbA1c levels through glycemic and/or non-glycemic pathways. Using reported association results for other glycemic traits, red blood cell counts (RBC), and iron traits, Chen et al [6] was able to classify 168 HbA1c-associated variants into glycemic (n=53), affecting iron level and/or iron metabolism (n=12), and regulating RBC traits (n=103). Besides HbA1c, based on GWAS Catalog, *HK1-rs17476364* was significantly associated with RBC and abnormal glucose (*P*<5e-8), *HK1-rs16926246* was significantly influencing hemoglobin (*P*<5e-8), whereas the remaining three SNPs (*GCK- rs2971670*, *GCK-rs4607517*, and *G6PC2- rs560887*) were known genetic determinants of fasting blood glucose (*P*<5e-8). In line with these reported findings, associations of two *HK1* SNPs with Hb levels, as well as associations of *GCK* SNPs and *G6PC2* SNPs with fasting blood glucose were validated in our analyses. Therefore, *HK1* gene influence HbA1c levels through non-glycemic pathways, and both *GCK* and *G6PC2* genes regulate HbA1c levels through glycemic pathways.

*HK1* gene (10q22.1) encodes a protein Hexokinase 1 enzyme, and acts as a priming step of glucose metabolism and ATP production by phosphorylating glucose to glucose-6-phosphate (G6P) [18]. Due to its absent expression in primary Glucose-Regulating cells (normal pancreatic β-cell and hepatocytes), *HK1* gene is not involved in the regulation of physiological blood glucose levels [19]. This explains why neither we nor others detected significant associations of *HK1* genetic variants with blood glucose levels in nondiabetic participants. Due to lack of mitochondria and oxidative phosphorylation, the lifespan of RBCs depends on a special version of *HK1* gene catalyzed ATP production through anaerobic glycolysis [20]. These molecular mechanisms provide a functional basis for the significant associations between *HK1* genetic variants and Hb levels reported by our group and other investigators, as well as how *HK1* gene influences HbA1c levels through non-glycemic pathways. Compared to *HK1*, the *GCK* gene (7p13) encodes Glucokinase, which catalyzes the same first step of glycolysis, but functions primarily in Glucose-Regulating cells, including pancreatic β-cells and hepatocytes [21]. While the *G6PC2* gene (2q31.1) encodes Islet-specific Glucose-6-Phosphatase (IGP), which also functions primarily in pancreatic β-cells, but catalyzes the dephosphorylation of glucose-6-phosphate back into free glucose [22]. Thus, *GCK* and *G6PC2* jointly set the β-cell glucose threshold for blood glucose control [23], ultimately affecting HbA1c levels through glycemic pathways. Based on the facts that *HK1* (10q22.1), *GCK* (7p13), and *G6PC2* (2q31.1) are located at different chromosomes, do not encode transcription factors, and exhibit different expression pattern, we can rule out the direct physical interactions between *HK1* and *GCK* genes, and between *HK1* and *G6PC2* genes. Since both HK1 activity and circulating glucose levels are key determinants of RBC lifespan, the significant interaction effects of *HK1-GCK* and *HK1-G6PC2* on Hb and HbA1c levels are likely driven by functional interplay between HK1-dependent anaerobic glycolysis and glucose regulation mediated by *GCK* and *G6PC2*.

This analysis is inherently imperfect. First, as our findings were obtained from participants of European ancestry, investigators should interpret the results with caution when applying them to the general population or to other ethnic groups. Second, due to the substantial computation demands of genome-wide epistatic analyses, we restricted our epistatic analyses to 133 known HbA1c-associated variants, and therefore interrogated only a small fraction of the human genomes. Third, our replication in FOS did not account for genetic PCs and family relatedness, which may have inflated the observed interaction effects. However, the consistent significance across FOS and HRS provides additional support for our findings. Fourth, the functional roles of five intronic variants in *HK1*, *GCK*, and *G6PC2* in regulating the expression of their respective genes remain unclear. Fifth, the exact molecular mechanisms underlying the significant interactions between *HK1* and *GCK* genes, as well as between *HK1* and *G6PC2* genes, remain unclear. Additional functional studies are needed to elucidate how these gene pairs jointly influence HbA1c.

In summary, we identified significant *HK1-GCK* and *HK1-G6PC2* interactions, yielding novel biological insights into HbA1c regulation. More broadly, gene-gene interactions and genome-wide epistasis analyses may further unravel the molecular architecture of complex traits, and improve PRS performance.

## CRediT authorship contribution statement

Lihua Wang: Writing – review & editing, Writing – original draft, Validation, Software, Methodology, Investigation, Formal analysis, Data curation, Conceptualization. Joseph H. Lee: Writing – review & editing, Supervision. Bharat Thyagarajan: Writing – review & editing, Supervision. Anatoliy I. Yashin: Writing – review & editing, Supervision. E. Warwick Daw: Writing – review & editing, Supervision. Thomas T. Perls: Writing – review & editing, Supervision. Kaare Christensen: Writing – review & editing, Supervision. Joseph M. Zmuda: Writing – review & editing, Supervision. Michael A Province: Writing – review & editing, Supervision, Funding acquisition. Ping An: Writing – review & editing, Writing – original draft, Validation, Software, Methodology, Investigation, Formal analysis, Data curation, Conceptualization.

## Data availability

LLFS clinical data and WGS data can be downloaded at ELITE Synapse portal (syn52663596). FOS data is available at dbGAP with accession phs000007.v35.p16 and phg002135, and HRS data can be downloaded at https://hrs.isr.umich.edu with accession phs000428.v2.p2.

## Acknowledgments

The authors thank E. Warwick Daw for his contribution to LLFS WGS data quality control and FOS data acquisition, and Mary K Wojczynski for HRS data acquisition. We are grateful to LeAnne Kniepkamp for her administration support.

The authors thank the participants and staff of the Framingham Heart Study and the Health and Retirement Study for their contributions. The Framingham Heart Study is conducted and supported by the National Heart, Lung, and Blood Institute (NHLBI; Contract No. HHSN268201500001I) in collaboration with Boston University (Contract No. N01-HC-25195). This manuscript was not prepared in collaboration with investigators of the Framingham Heart Study and does not necessarily reflect the opinions or views of the Framingham Heart Study, Boston University, or NHLBI.

The Health and Retirement Study is sponsored by the National Institute of Aging (grant number NIA U01AG009740) and is conducted by the University of Michigan.

## Abbreviations

HbA1c: Hemoglobin A1c
T2D: type 2 diabetes
LLFS: Long Life Family Study
BMI: Body mass index
PCs: Principal components
BIMBAM: Bayesian Imputation Based Association Mapping
BF: Bayes Factor
PRS: Polygenic Risk Score
HPLC: high-performance liquid chromatography
NGSP: National Glycohemoglobin Standardization Program
WGS: whole genome sequencing data
MGI: McDonnell Genome Institute
HWE: Hardy-Weinberg equilibrium
FOS: Framingham Heart Study Offspring Cohort
HRS: Health and Retirement Study
CIDR: Center for Inherited Disease Research
NHANES: National Health and Nutrition Examination Survey
RBC: Red blood cell
Hb: Hemoglobin

